# Impact of implementation of rapid syndromic molecular diagnostics on self-reported clinical and public health practice: a qualitative study in small island health services

**DOI:** 10.1101/2024.11.05.24316772

**Authors:** Riinu Pae, Adam Millest, Anna Tirion, Matthew Dryden, John E. Lee, Natalie Wight, Graham Fraser, Janice Toplass, Dale Weston

## Abstract

**Objective:** This evaluation aimed to assess the impact of implementing Biofire© filmarray rapid syndromic molecular diagnostics systems in UK Overseas Territories, which are small jurisdictions with historically limited microbiological diagnostic capacity. The diagnostic systems were installed to improve patient management and public health response.

**Methods:** We conducted a qualitative evaluation. Data were gathered through semi-structured interviews and a focus group with clinicians, laboratory staff, and Chief Medical Officers.

**Results:** The interviewees’ reported substantial improvements in diagnostic capabilities. Interviewees shared that implementation reduced test turnaround times to 1-24 hours compared to several days to weeks pre-implementation, enabling faster and more accurate clinical management and improving clinician and patient satisfaction. Reduced reliance on off-island reference laboratories and patient medical evacuations was reported, contributing to potential cost savings and increased health system resilience. Respondents found rapid diagnostics to be useful in the context of communicable disease outbreaks. However, high test cartridge costs, supply and logistics issues, and lack of or low utilisation of protocols were noted challenges.

**Conclusion:** The implementation of this rapid automated syndromic molecular diagnostics technology markedly enhanced diagnostic capacity in territories included in this evaluation, particularly for respiratory, bloodstream, and gastrointestinal infections. This advancement accelerated diagnosis, was seen to improve patient management and antimicrobial stewardship. Despite these benefits, challenges remain. Further research is needed to assess the long-term impact on clinical practice, health outcomes, and cost-effectiveness, particularly in the unique contexts of small islands.

## INTRODUCTION

Small, resource-limited health jurisdictions, such as the Overseas Territories of the United Kingdom (UKOTs), have historically faced significant challenges in maintaining effective microbiological diagnostic services. These challenges stem from low test volumes, limited available tests, and logistical difficulties in referring samples to regional laboratories, and result in delays in the identification and management of infections. These factors can negatively affect clinical management, antimicrobial resistance assessments, public health surveillance and outbreak response. The challenge of provision of modern laboratory medicine to low-income countries has been recognised as a global challenge (1).

The COVID-19 pandemic exacerbated issues such as limited laboratory capacity, difficulties recruiting and retaining specialist trained staff, and challenges ensuring adequate supply of consumables and equipment, leading to delayed diagnostic results. In response to these vulnerabilities, the UK Health Security Agency (UKHSA) UKOTs Public Health Programme, funded by the Foreign and Commonwealth Development Office, successfully supported the territories in implementing open platform reverse transcription polymerase chain reaction (PCR) technology for COVID-19 diagnosis. However, maintaining PCR testing proved difficult due to workforce skills gaps and capacity limitations. UKHSA’s UKOT Laboratory Strengthening Project aimed to address these limitations through the use of rapid, automated syndromic molecular diagnostics, deploying closed molecular assays – GeneXpert and FilmArray Biofire technology – to provide rapid syndromic testing for a broad range of pathogens. The *Biofire© filmarray*© panels enable rapid detection of gastrointestinal, respiratory, bloodstream, and neurological infections, including antibiotic resistance genes. This technology delivers differential diagnosis for single samples within an hour and is particularly well-suited to small laboratories with low test volumes, offering an effective solution to enhance local diagnostic capacity and reduce dependence on reference laboratories.

In March 2023, UKHSA funded and delivered *Biofire* equipment and reagents to eight UKOT laboratories, including Montserrat, Anguilla, the British Virgin Islands, the Turks and Caicos Islands, the Cayman Islands, St Helena, the Falkland Islands, and Ascension. Other territories, such as Bermuda and Gibraltar, had already developed molecular diagnostic capacity independently, while very small territories like Pitcairn Island and Tristan da Cunha were not included as they lack diagnostic laboratories. The system was implemented between May and June 2023, supported by on-site visits, online training, and awareness-raising events within health professionals’ networks (detailed in a sister paper describing the implementation and clinical impact of implementation (2)).

The introduction of the technology was expected to achieve several key outcomes: improved clinical management, enhanced infection surveillance, better outbreak management, and more effective control of antibiotic resistance. However, successful implementation requires integration into existing laboratory services, uptake and optimal usage by clinicians and laboratory staff. Given the broad range of factors that influence the adoption of new healthcare processes and clinician investigation practices (3–5), it is essential to understand how the *Biofire* system was implemented, what changes – if any – were observed, and how its usage could be optimised.

This paper evaluates the clinical implications and impact of *Biofire* implementation across eight UKOTs, offering insights into the integration and effectiveness of this technology in enhancing diagnostic capabilities within these small, resource-challenged territories. The evaluation focused on three main questions:

1. How was the technology used in its first year of implementation?
2. What impact has the installation of the technology had?
3. How could the usage of this technology be optimised?

## METHODS

### Design

The initial evaluation design was mixed-methods with a pre-peri-post questionnaire, focus groups 1-year post-implementation and quantitative laboratory data. However, as the questionnaire response rate was low, despite continued promotion of the evaluation in relevant staff networks, we shifted to a qualitative evaluation.

Quantitative laboratory data is reported in (2), a sister paper describing the clinical impact of implementation.

### Participants

Nine participants from six of the eight UKOTs where RASM has been implemented shared their views. This included five clinicians and four Chief Medical Officers (CMO), who are medical doctors serving as the top government advisors on health. Three of these islands (Anguilla, the Cayman Islands, Montserrat) are in the Caribbean region, and three (Ascension Island, the Falkland Islands, St Helena) are in the South Atlantic. Four clinicians (three doctors, one Biomedical Scientist) and one CMO were interviewed individually, while a focus group included three CMOs and one Laboratory Manager. Additionally, three UKHSA representatives responsible for implementing the system (MD, JT, JL) participated in the focus group for informational support.

### Data collection and sampling

A researcher from the UKHSA UKOTs Programme (RP) conducted five semi-structured interviews. Participants were recruited through UKOTs health professionals’ networks using snowball sampling. The interviews, conducted online via Microsoft Teams, ranged from 6 to 33 minutes (*M* = 22) and took place in December 2023 and January 2024.

Focus group participants were recruited using convenience sampling through the UKOTs CMO network. A regularly scheduled virtual meeting was used to hold a 45-minute focus group in April 2024 via Microsoft Teams, facilitated by a UKHSA researcher DW.

### Procedure

Interview and focus group participants were introduced to the evaluation’s purpose in the invitation email and at the start of the discussion. All were asked for verbal consent. Focus group participants were informed that if they did not wish to participate or answer specific questions, they could refrain from doing so by not speaking. The topic guides focused on experiences requesting and conducting testing before and after the installation of RASM, what changes – if any – they had observed, and how to optimise its usage (Supplementary materials 4, 5 and 6).

### Qualitative data analysis

The focus group discussion and interviews were audio-recorded and transcribed using Microsoft Teams, then checked for accuracy and pseudonymised. Transcripts were imported into NVivo 14 for inductive thematic analysis, conducted by AT (6). Responses were coded and organised into groups based on research questions, iteratively refining codes and themes. As the focus group was conducted after we had analysed the interviews, existing codes from interviews were applied to the focus group discussion, with new codes cross-referenced and themes expanded or newly generated as appropriate. Distinctions were made where themes or subthemes applied only to CMOs or clinical staff, though experiences were generally similar due to CMOs’ clinical backgrounds. Supplementary material 6 contains the complete codebook, including example quotes illustrating each code.

### Ethics

The UKHSA Research Ethics Governance Group (REGG) granted ethical approval for this evaluation (NR0374), including the modification of the design.

## RESULTS

The key findings are summarised in Table 1.

**Table 1.**
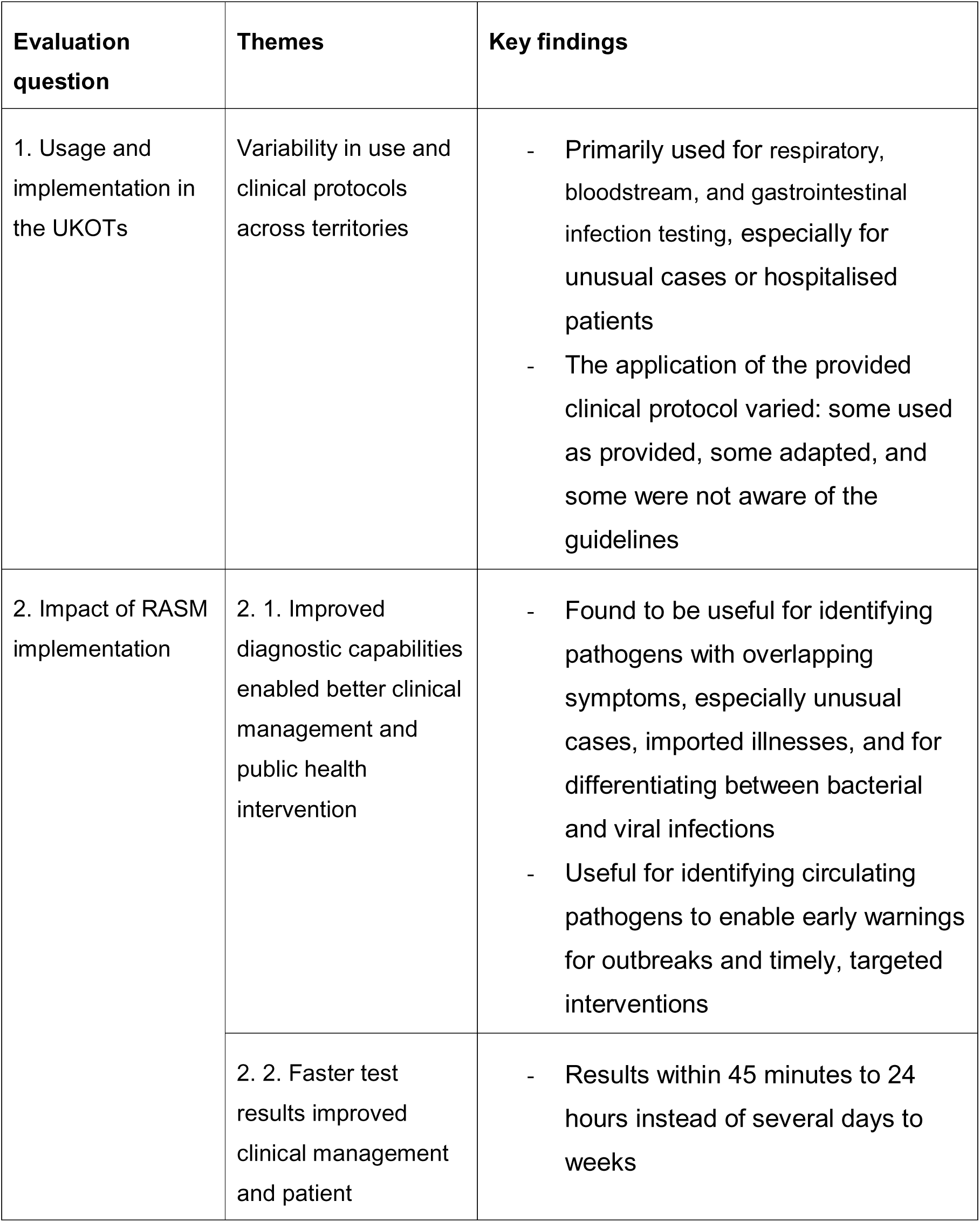
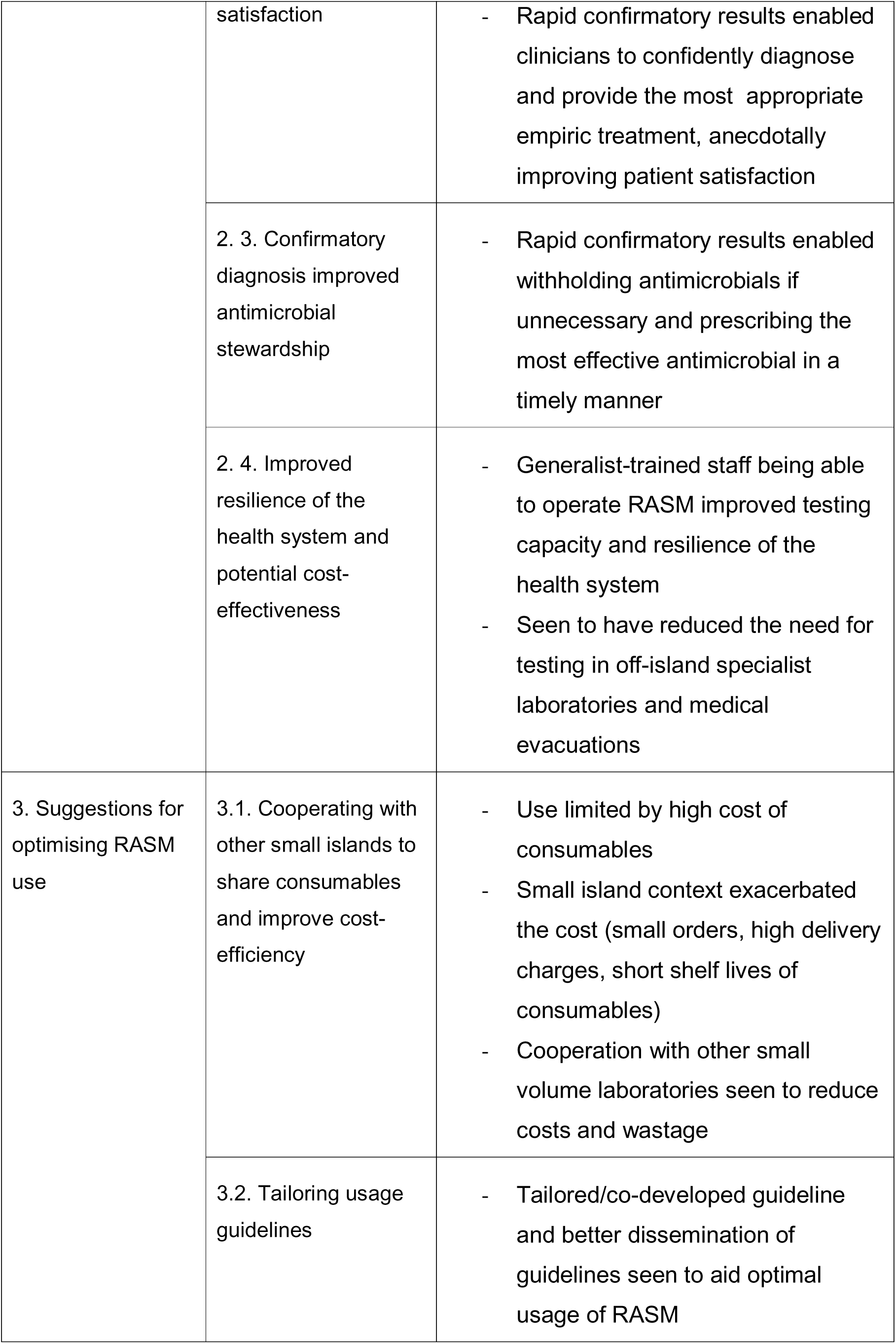
Key findings.

### 1. Usage and implementation in the UK Overseas Territories

In the first year, RASM was primarily used for respiratory, bloodstream, and gastrointestinal infection testing. Bloodstream infections were prioritised due to their severity and the need for faster results in individual patient management, while the gastrointestinal panel was reserved for clusters, outbreaks and serious individual cases to support public health and patient management, and to minimise costs when cheaper, more traditional laboratory diagnostic methods could be used instead. RASM was often limited to hospitalised patients, unusual cases, or situations where the result would directly influence patient management. CMO 4 summarised this approach: “*We only do the Biofire here on people who have significant symptoms and where a diagnosis will either alter their individual management or may have public health implications if there’s a viral outbreak.”.* Neurological testing was not mentioned at all in the interviews, likely due to the low incidence of neurological infections.

To support implementation, UKHSA technical experts provided training (MD) and written protocols including clinical criteria for testing (MD, GF, NW) to guide appropriate clinical use of the RASM (Supplementary Material 2). However, the implementation of these guidelines varied. Some participants used the guidelines as provided, some adapted it locally by applying illness severity or hospitalisation as criteria for testing. Others relied on case-by-case judgement, either because they were unaware of the guidance or chose not to apply it. For example, Doctor 2 noted, “*We don’t have a protocol set up. We take it as an individual and we discuss the patients (…). You just have to use your own discretion.*”.

Decision-making processes also varied. In one small territory, the CMO was directly involved in each testing decision due to their hands-on role in clinical work. In others, doctors or laboratory scientists requested RASM tests independently, consulting or informing the CMO only when necessary.

### 2. Impact of RASM implementation

#### 2.1. Improved diagnostic capabilities enabled better clinical management and public health intervention

Before the implementation of RASM, some UKOTs had limited or no capacity to detect pathogens, particularly for respiratory and enteric illnesses. Clinicians often had to conduct multiple tests to identify the microbiological cause of the illness or conduct no tests at all. Following *Biofire* implementation, they were able to detect a greatly expanded range of pathogens in all syndromes covered.

Clinicians found RASM especially useful for identifying pathogens in patients with overlapping symptoms, particularly in unusual cases, imported illnesses, or infections requiring specific treatment. Doctor 3 explained: *“When you’re in a tropical space like ours, where there are multiple viruses that can present in similar ways and with varying management, it’s always helpful to be a little bit more definitive and to have that final answer”*.

More broadly, RASM enabled better differentiation between viral and bacterial infections, improving treatment decisions and antimicrobial stewardship. As Doctor 1 described: *“The main thing for me as a clinician in the frontline is to actually have a quick answer to one question: is this a viral or bacterial infection? And also what I found very useful is to have an indication about what the bacteria might be in the blood.”*.

Clinicians also reported that RASM was valuable in offering early warnings for outbreaks, identifying circulating pathogens, and enabling timely, targeted interventions. Doctor 2 explained how they were able to provide more accurate information to the community during a potential outbreak: *“(we said in the radio): ‘Look, we’ve tested a few people. These are the viruses going around. It’s not COVID. You’re not gonna die, and it’s probably gonna last this long. It’s this virus. Just take paracetamol, fluids and don’t overrun the clinic. You don’t need to see the doctor.’ ”.* This was seen to improve patient satisfaction and reduce further testing when symptoms aligned with known circulating infections.

#### 2.2. Faster test results improved clinical management and patient satisfaction

Participants reported that before RASM, test results often took several days to weeks due to the methods used, the need for specialist staff with limited availability during out-of-hours, and the need to send samples to off-island reference laboratories. A Biomedical Scientist explained: *“Culture obviously would have been like 2 days at least for plates to get incubated, and any positives from that would maybe take another 40-48 hours (for identification and antibiotic sensitivities).”.* In contrast, RASM provided results within 45-60 minutes for organism identification and resistance mechanism detection. Participants noted that test results were typically available within 24 hours of patient admission, with urgent cases receiving results in 1-3 hours —particularly valuable for treating severely ill patients.

Clinicians reported that RASM has had a positive impact on both their and patients’ satisfaction by providing rapid confirmatory results, enabling the clinician to confidently diagnose, provide the most appropriate treatment, and inform patients of the likely trajectory of the disease.

#### 2.3. Confirmatory diagnosis improved antimicrobial stewardship

The rapid confirmatory results enabled clinicians to prescribe the most effective antimicrobial in a timely manner or withhold antimicrobials if shown to be unnecessary. In contrast, clinicians reported that prior to RASM they sometimes felt “stuck” or “pressed into a corner” and prescribed antibiotics out of caution or as a temporary measure while waiting for test results.

#### 2.4. Improved resilience of the health system and potential cost efficiency

Participants praised RASM for its ease of use, noting that, unlike traditional plate/disk diffusion testing methods, it can be operated by generalist-trained staff such as clinicians and laboratory assistants. This was especially valuable in territories with small medical workforces, as it improved testing capacity and resilience without requiring specialised microbiology training. A Laboratory Manager explained: *“[RASM] has definitely improved our workflow, our resilience. Specifically, if a microbiologist, for whatever reason, is sick, we don’t always have two people here on island that can do the intricate micro testing like blood cultures, sputums and the like. And the [RASM] has revolutionised that aspect for us and given us resilience which we didn’t have before.”*

A Biomedical Scientist from another territory shared the view: *“Microbiology, it’s a lot of experience and training to learn to identify organisms in plates and then there’s a lot of manual manipulation of organisms. So if somebody doesn’t have that background, they can just set up the [RASM] because it gives a result straight away.”*

RASM enabled a wider range of tests to be conducted locally, reducing— but not eliminating —the need for off-island testing and contributing to faster result times overall. Participants suggested that the fast, accurate results of RASM might offset its costs by reducing the need for multiple tests and allowing for more timely treatment, which minimises the burden on the health system. Medical evacuations to receive urgent care abroad, which are particularly expensive, were also reduced. The Laboratory Manager described: *“[RASM] has also cut down potential med-evacs that we may not have needed. Specifically, during our tourist and fishing seasons, where we have people come from all around the world, which exposes us to diseases that aren’t normally found in this part of the world. We see weird and wonderful things pop up just for maybe a couple of weeks of the year, and being able to diagnose things or rule things out, which is just as useful, without having to spend copious amounts of money flying them out to [closest continent] has been a great stress relief for the rest of us.”*

Additionally, a CMO of a tourism-dependent island argued that the income generated by proving the island’s safety through RASM testing helps offset its costs.

### 3. Recommendations to optimise RASM use

#### 3.1. Cooperating with other small islands to share consumables and improve cost-efficiency

While RASM was considered an extremely valuable asset, the high cost of £120 per test cartridge restricted its use. CMO 4 stated: *“[RASM] is probably not something we would have purchased because we have a limited budget and (…) it doesn’t get used very often. But having said that, it’s very valuable to have it here.”* The Laboratory Manager explained how the specific context of a small island further increased the cost: *“It’s expensive per test, magnitudes more expensive than plates and with expiry dates that are only several months long which can be quite good for certain areas, but for us, when we’re having to buy things in bulk and a couple of the items being classed as hazardous, so we have to pay additional cost for that hazardous rate to get to us, yeah, the cost per test is quite large.”*

The high cost was exacerbated by short shelf lives of the consumables and strict expiry dates after which the machine would reject the panels, leading to wastage of expensive consumables. This was a particular challenge for the smaller UKOTs with low testing volumes: *“For us, it’s all about the economy of scale. (…) We’re liable to lose most of the cartridges we purchase initially because of the expiry.”* (CMO 2), and for outbreak preparedness: *“You’re always caught by what you need to keep in stock. Particularly if you’re going to be in an outbreak situation and you’ve got to increase the numbers of people that you’re looking at and batching and pooling of specimens and having that pre thought out for all of your diagnostic system.”* (CMO 3).

To address this, several participants, particularly CMOs, suggested cooperating with other territories in the region to reduce costs and wastage by ordering consumables in bulk and dividing them as needed. A participant proposed coordinating shipments along shared supply routes, stating: *“There is a flight that goes from the UK to the [Territory 2], via [Territory 1] and the flight that goes from [Territory 1] to [Territory 4]. If we bulk bought stuff which would cut down the amount of cost that we’re having to fish out basically, and divvy it out amongst people along that supply route. We’d cut down A) the cost per island and B) probably utilise it better before it runs out and shelf life because we have to end up buying stuff in bigger bulk than we necessarily would need just so we can justify the cost of the freight.”*

Alternatively, respondents suggested that subsidies may help make RASM more accessible in low-resource territories.

#### 3.2. Tailoring usage guidelines

Although one clinician felt that the small population and close-knit community of clinicians negated the need for a written protocol, most participants thought that it would be useful to have a concrete and locally tailored protocol detailing when RASM should and should not be used and why. This was seen to prevent subjective decision-making and ensure effective use of the system. Although training and guidelines were provided upon installation (Supplementary Material 1), the intended audience were either unaware of these guidelines or did not apply them in practice as intended, necessitating a better tailoring and dissemination of the guidelines.

## DISCUSSION

This is one of the few evaluations of a rapid automated syndromic molecular (RASM) diagnostics technology *Biofire FilmArray* in the unique context of six UK Overseas Territories (UKOTs). These small island territories, some of which are small island developing states (SIDS), face unique challenges due to their limited healthcare infrastructure and diagnostic capabilities. Notably, this evaluation took a qualitative approach, a rarity in diagnostic technology evaluations, complementing a sister paper detailing the implementation and clinical impact of RASM (2). A range of perspectives from key stakeholders, such as Chief Medical Officers, clinicians, and laboratory staff, provided a holistic understanding of RASM’s impact across different levels of the healthcare system.

The implementation of RASM reportedly enhanced diagnostic capabilities, particularly for respiratory, bloodstream, and gastrointestinal infections, with clinicians observing marked reductions in test turnaround times from multiple days to an hour. This accelerated diagnosis was perceived as a critical improvement in patient management and public health interventions, allowing for more timely and targeted treatments, thereby supporting antimicrobial stewardship (AMS).

The unique context of SIDS magnifies both the benefits and challenges of RASM implementation. Small islands have a limited ability to treat complex or rare conditions on-island, necessitating costly medical evacuations and sample transport to specialist laboratories. As the confirmatory diagnoses provided by RASM enabled managing unusual cases with more certainty and confidence, it was seen to have reduced medical evacuations and the need to send samples off-island. The system can be operated by generalist-trained staff, so the implementation was seen to enhance the resilience of these small healthcare systems, which often struggle with attracting and retaining highly trained microbiology staff.

Despite the benefits in small island contexts, the broader literature on the impact of RASM diagnostics is mixed (e.g. (7–10). Most previous evaluations with mixed impact have been conducted in high-resource settings with preexisting advanced capacity and capabilities. However, even modest improvements in diagnostic capacity have had profound impacts on patient care in low-resource settings with previously minimal or non-existent diagnostic capabilities, like territories in the study (11, 12). This suggests that the impact of RASM depends on the existing diagnostic capacity and infrastructure (13).

Furthermore, literature regularly shows that the implementation of diagnostic systems alone is insufficient to change clinical practice (14, 15). Inconsistent protocol adherence, misinterpretation of results, and a lack of follow-through on diagnostic findings can undermine the potential benefits (1, 13, 16). Therefore, successful implementation requires more than just providing equipment and training; it necessitates addressing behavioural, organisational, and contextual factors that hinder or support the innovation’s optimal use (5). Our study found that while guidance had been produced and disseminated, it was applied inconsistently, underscoring the importance of engaging stakeholders in the co-development of implementation strategies and guidance to foster a sense of ownership and ensure tailoring to the local health system’s needs (14, 16).

Another critical consideration is the role of rapid diagnostics in AMS. RASM technologies have shown promise in AMS, however, the impact has been mixed (10, 17, 18). While one study found no additional benefits of integrating an AMS guidance with RASM diagnostics (18), others emphasise that AMS decision-making support is essential to fully realise the potential of these technologies (10, 16, 19, 20). The effectiveness of these systems in AMS depends heavily on accurate result interpretation and access to follow-up care: without AMS support, the risk of overtreatment due to false positives and challenges in interpreting co-infections remains high (10, 11). Thus, integrating AMS programmes with RASM is crucial for optimising their benefits and avoiding overtreatment.

There is also emerging evidence, including from this evaluation, finding RASM potentially beneficial for outbreak identification and management (21). However, as with AMS, broader factors such as lack of capacity and capabilities for outbreak investigations and public health action in small island contexts may limit its impact. Therefore, impact of RASM on outbreak management requires further evaluation.

This evaluation contributes to the literature by highlighting the impact of RASM on clinician and patient satisfaction, an area overlooked in previous research. Compared to the pre-implementation period, when diagnostic capabilities were limited or testing was slower, the ability to obtain results quickly led to improved satisfaction among clinicians and, anecdotally, among patients. While some studies touch on satisfaction (e.g., (15)), there is a paucity of social research examining how the implementation of new health technologies, particularly microbiology diagnostic systems, affects the satisfaction of those using or benefiting from them. Future studies should adopt a more holistic approach to evaluating diagnostic innovations, including user experiences and satisfaction alongside clinical outcomes and cost-effectiveness.

The cost of purchasing and running these systems is a common barrier for low-resource settings – including ours, but this was further compounded by the low testing volumes and complex supply chains unique to small island contexts. The cost, together with logistical and supply chain issues, can limit their impact due to inconsistent availability or underuse (11, 16, 22). Moreover, the reliance on external support – the diagnostics were funded by the Foreign and Commonwealth Development Office and the implementation supported by the UKHSA UKOTs Laboratory Strengthening Programme – raises questions about long-term sustainability. Once the funding ends, the costs of maintaining these systems could become prohibitive, potentially limiting their use over time (1). While cooperating with other territories to bulk-buy consumables may improve the cost-efficiency of RASM, further research is needed on planning and managing laboratory stock in low-volume laboratories, the long-term cost-effectiveness, and medium- and long-term impacts of this technology in SIDS. Finally, studies could also explore alternative funding models and partnerships to ensure sustainability beyond the initial grant periods for territories that need these (such as those eligible for Official Development Assistance).

Despite the broader findings and implications relevant to small islands, there are also limitations to consider. We faced challenges in recruiting for the questionnaire study, necessitating changing the research design to prioritise in-depth insights from a smaller sample. As a result, it is possible that the participants with positive experiences with RASM were overrepresented, while those with neutral or negative views may have been underrepresented, introducing a potential bias in the findings. Furthermore, the healthcare infrastructures and resources of the territories vary widely, which may limit the generalisability of the results to other low-resource settings. Despite these limitations, this evaluation captured a diverse set of views from three-quarters of the territories where RASM was implemented, representing a reasonable proportion of the small health workforces, providing novel insight into a previously underexplored area.

## CONCLUSION

Overall, this study provides important insights into the implementation of advanced diagnostic technologies in small island contexts, particularly highlighting how RASM technology significantly enhanced diagnostic capacity for respiratory, bloodstream, and gastrointestinal infections in eight UKOTs—a crucial advancement for islands with previously limited diagnostic capabilities. However, further studies are needed to fully understand its impact on clinical practice, health outcomes, and long-term cost-effectiveness, particularly in resource-limited settings.

## Data Availability

All data produced in the present study are available upon reasonable request to the authors.

## DECLARATIONS

## Acknowledgements

The authors would like to thank the interview and focus group participants. Without their contributions, this work would not have been possible.

## Ethical approval

The Research Ethics Governance Group at UK Health Security Agency granted ethical approval for this evaluation (NR0374), including the modification of the design.

## Authors contribution

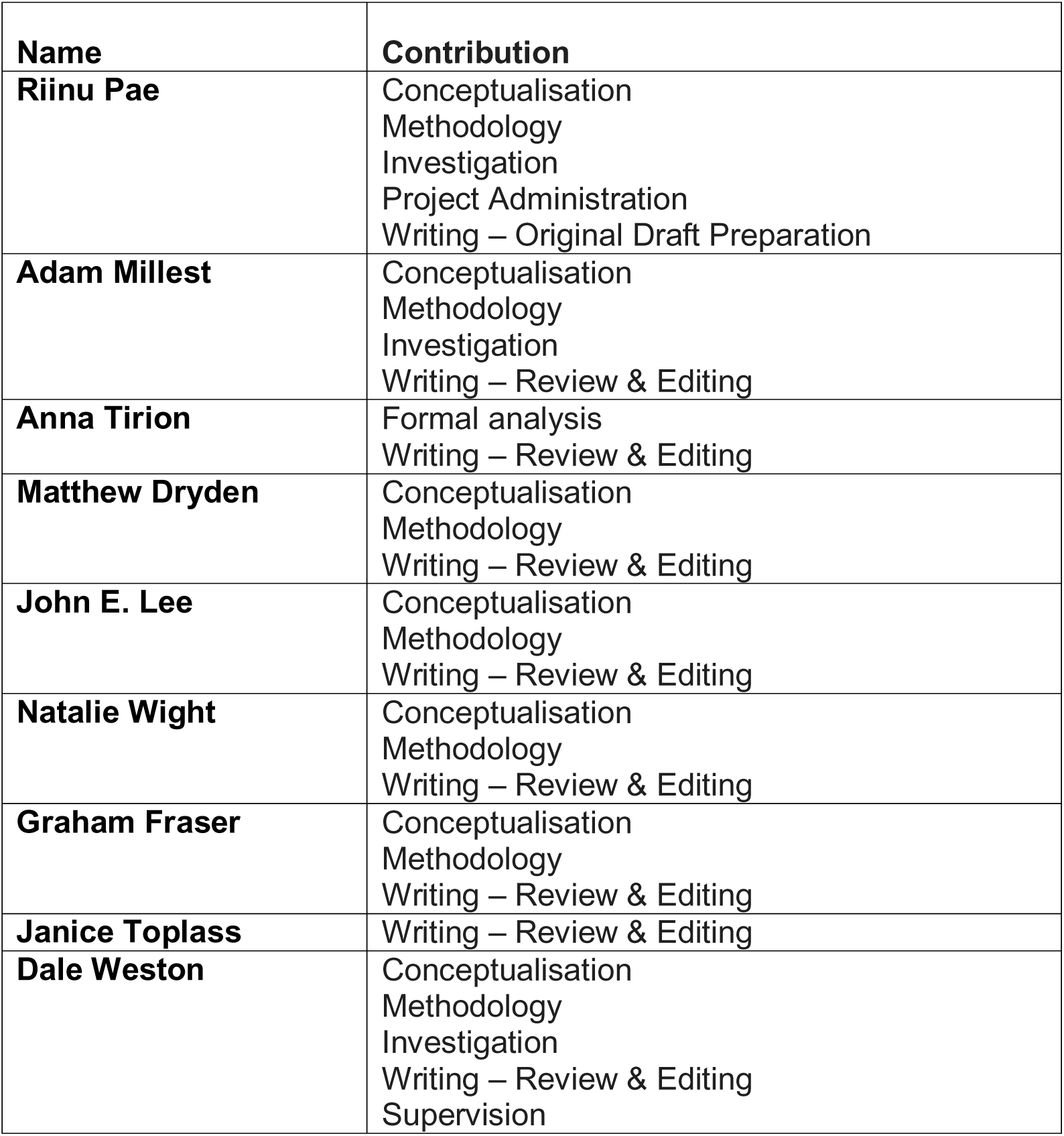

## SUPPLEMENTARY MATERIALS

Not uploaded. Available upon reasonable request to the authors.

